# COVID-19 and heart medications: What’s the connection?

**DOI:** 10.1101/2020.08.24.20174367

**Authors:** Nafiseh Saleknezhad, Bardia Khosravi, Amir Anushiravani, Masoud Eslahi, Amir Reza Radmard, Azin Sirusbakht, Seyed Mohammad Pourabbas, Mohammad Abdollahi, Amir Kasaeian, Majid Sorouri, Ali Reza Sima

**Affiliations:** Digestive Disease Research center, Digestive Disease Research Institute, Tehran University of Medical Sciences, Tehran, Iran; Department of Internal medicine, Shariati hospital, Tehran university of medical sciences, Tehran, Iran; Department of Radiology, Shariati Hospital, Tehran University of Medical Sciences, Tehran, Iran; Hematology, Oncology and Stem Cell Transplantation Research Center, Tehran University of Medical Sciences, Tehran, Iran

**Keywords:** COVID-19, Hypertension, Ischemic Heart Disease, SARS-CoV-2, Statins, Treatment

## Abstract

**Aim:** To determine association between clinical outcome of COVID-19 and prior usage of cardiovascular and metabolic drugs, including, Aspirin, ACEIs, ARBs, Clopidogrel, metformin, and Statins.

**Methods:** Statistical examination of the demographic, clinical, laboratory and imaging features of 353 patients with SARS-CoV-2 disease admitted from February to April 2020.

**Result:** Minor discrepancies were observed in the clinical presentations, radiologic involvement and laboratory results across groups of patients under treatment with specific drugs. Aspirin-users had better clinical outcome with lower need of ventilation support, whereas, metformin- users had increased chance of intubation and of mortality.

**Conclusion:** Although not being conclusive, our findings suggest the possibility of the effect of previous drug usages on the various presentations and clinical course of COVID-19 infection.

## Introduction

Late 2019, is marked by the emergence of a new type of beta-coronavirus, which soon led to a disastrous outbreak of the so-called SARS-CoV-2 disease [1]. The outbreak was coined as a global pandemic by WHO in March 2020 after countries world-wide got involved [2]. By May 2020, more than four million known cases of the disease and about three hundred thousand of deaths were reported globally by the 116^th^ WHO situation report.

Due to the notable death toll of SARS-CoV-2, risk factors that predispose patients to mortality and more severe forms of the disease are under investigation. It has been well observed that individuals burdened with hypertension, diabetes or ischemic heart disease are at higher risk of developing severe manifestations of SARS-CoV-2, or of mortality due to the infection [3-5]. Although it is reasonable to maintain drugs that high-risk patients take due to their chronic medical conditions, there are certain considerations about their safety during COVID-19 infection. For instance, statins and angiotensin receptor blockers (ARBs) may actually worsen the outcome of COVID-19 patients, who, due to their underlying diseases, have poorer prognosis [6]. On the other hand, certain drugs that are used in similar conditions are thought to have a protective effect against the novel coronavirus. For example, metformin has been suggested to be added as an adjuvant therapy to diabetic, old, and even obese patients’ treatment for the sake of better outcome in COVID-19 setting [7].

Therefore, we conducted a study based on our registry of COVID-19 patients to evaluate the effect of those drugs, which are widely used in chronic diseases such as hypertension, diabetes mellitus and ischemic heart disease, on the clinical outcome and mortality rate of the patients inflicted with the novel coronavirus.

## Materials & Methods

### Study Design and participants

In a retrospective cross-sectional study at Shariati Hospital in Tehran, Iran, we included all patients admitted with the diagnosis of COVID-19 based on the criteria published by WHO on January 12th, 2020 [8], from February 25th, 2020, to April 21st, 2020. Patients with respiratory symptoms and one of the following were admitted: 1) loss of consciousness, 2) Respiratory rate more than 24, 3) pulse rate more than 90, 4) Systolic blood pressure less than 90 mmHg, 5) abnormal respiratory sounds, or 6) O2 saturation less than 93%. Furthermore, high-risk patients with respiratory symptoms or fever underwent computed tomography scans of the lung (CT scans). In case of a suggestive lung CT scan of COVID-19 infection, the patients were transferred to the COVID ward. Patients with a negative lung CT scan underwent further workup. Patients who were already admitted to the hospital were transferred to the COVID ward if they experienced COVID-19 infection during hospitalization. We used the census sampling method, and after exclusion of cases with missed data, four hundred and four patients were selected. Two radiologists reviewed all the images and patients with not suggestive lung CT scan and negative SARS-CoV2 RT-PCR were excluded. We extracted clinical data, laboratory findings, and lung CT scans from the remaining three hundred fifty-four patients’ medical records. The study was approved by the ethics committee of Tehran University of Medical Sciences. The ethics committee waived the requirement for informed patient consent for this retrospective study subject to the anonymity of patients.

### Measurements

The variables recorded are given in Table 1.

**Table 1:** Variables recorded and their categorization.

|  |
| --- |
| **History and physical examination:** |
| Age: young adults (<50 years old), middle-age (50-65 years old), and elderly (>65 years old) |
| Sex: Male, Female |
| Body mass index (BMI): severely obese (BMI>35) and not severely obese (BMI<35) |
| Temp (T): normal (< 37.8°C), low-grade fever (39°C-37.8°C), and high-grade fever (>39°C) |
| Systolic BP (SBP): low (<100mmHg), normal (100-140mmHg), and high (>140mmHg) |
| Diastolic blood pressure (DBP): low (<70mmHg), normal (70-90mmHg), and high (>90 mmHg) |
| respiratory rate (RR): normal (<25 per minute), and abnormal (>25 per minute), |
| pulse rate (PR): normal (<100 per minute) and abnormal (>100 per minute) |
| arterial O<sub>2</sub> saturation (SO<sub>2</sub>): normal (>95%), borderline (90%-95%), and low (<90%) |
| Presence or absence of:<br>chills, sore throat, weakness, anorexia, anosmia, chest pain, loss of consciousness |
| Type of supplementary oxygenations. |
| **Past Medical History:** |
| Diabetes (DM) |
| hypertension (HTN) |
| ischemic heart disease (IHD) |
| Chronic kidney disease (CKD) |
| cancer |
| organ transplant |
| **Laboratory Findings:** |
| complete blood cell count (CBC), |
| Ratio of circulating neutrophil to lymphocyte count (NLR) |
| C-reactive protein (CRP) |
| Creatinine |
| Liver enzymes: AST, ALT, Alkaline phosphatase (ALK-p) |
| Total bilirubin |
| Coagulation profile: Partial Thromboplastin Time (PTT), international normalized ratio (INR) |
| Venous Blood Gas: pH, PCO<sub>2</sub>, HCO<sub>3</sub>, |
| Urinalysis: hematuria (RBC >3 in hpf), leukocyturia (WBC>5 in hpf), proteinuria |
| COVID-19 RT-PCR |
| **Lung CT Scan:** |
| ground-glass opacity |
| consolidation |
| bilateral involvement |

The q-SOFA score was calculated for all patients, and a patient with q-SOFA Score ≥2 was categorized as high risk [9].

Outcomes included days of hospital stay, intubation, ICU admission, and in-hospital death.

### Statistical analysis

Categorical variables were described in frequencies and percentages. Quantitative variables were described with the mean (Standard deviation (SD)) or median and [Q1 - Q3]. Parametric and nonparametric tests, including t-test and Mann-Whitney test for comparing quantitative variables and Chi-squared test for comparing categorical variables were used. In a first step, variables showing associations at a significance level of α=0.20 in a univariable logistic models were selected for inclusion in the multivariable logistic model for adjustment. Statistical analyses were performed using Stata (Corp. 2009. Stata Statistical Software: Release 11. College Station, TX: StataCorp LP.). Statistical significance was defined as P < 0.05.

## Results

### General Findings

Our study was based on clinical, laboratory, and imaging data of 353 patients admitted with COVID-19. The majority of patients were males, with 203 (57.51%) cases out of 353. The distribution of patients in different age groups showed an increase in prevalence with respect to age, with patients older than 65-years compromising 45% of all cases. There were 21 (7.87%) severely obese patients with body mass index (BMI) >35.

With respect to the past medical history of cases, there were 129 (36.54%) patients with hypertension, 111 (31.44%) with history of diabetes, 91 (25.78%) with ischemic heart disease, and 75 (21.25%) individuals with no notable underlying disease. Less frequent medical conditions were cancer (45, 12.75%), hyperlipidaemia (18, 5.10%), thyroid disorders (16, 4.53%), organ transplantation (9, 2.55%), history of seizure (3, 0.85%), and multiple sclerosis (3, 0.85%). There was no HIV-positive patient recorded in our dataset.

At the two extremes of the spectrum of the clinical manifestations, it is noteworthy that 10 (2.83%) patients were found to be clinically asymptomatic, whereas 31 (8.78%) cases had a decreased level of consciousness upon admission. Considering the cardinal signs and symptoms of the disease, there were 198 (56.09%) cases with the subjective report of dyspnea, 178 (50.42%) with fever, and 177 (50.14%) with coughing. Other recorded manifestations of the disease were weakness (145, 41.08%), tachycardia (127, 37.03%), myalgia (65, 18.41%) nausea (44, 12.46%), tachypnea (43, 12.46%), anorexia (39, 11.05%), chest pain (26, 7.37%), chills (23, 6.52%), diarrhoea (18, 5.10%), sore throat (16, 4.53%), headache (15, 4.25%), coryza (8, 2.27%), and anosmia (1, 0.28%). Here, we can note more specific clinical findings. From the total number of 348 patients with recorded core body temperature, there were 94 (27.01%) and 17 (4.89%) cases with 37.8-39 °C and >39 °C respectively. Majority of patients (154, 44.27%) had borderline blood oxygenation (90-96%) measured by pulse oximetry, while 132 cases (37.93%) had normal and 62 cases (17.82%) had low blood oxygenation, out of 348 total recorded cases. Both systolic and diastolic blood pressures in the most patients were found to be in normal range upon admission.

The most notable laboratory findings were abnormal serum aspartate aminotransferase (AST) levels (50.56%; 136 out of 269), positive C-reactive protein (CRP) (50%; 157 out of 314), lymphopenia (44.44%, 108 out of 243), abnormal serum alkaline phosphatase (Alk-p) level (41.83%; 110 out of 265) thrombocytopenia (31.33%; 104 out of 332), proteinuria (38.55%; 64 out of 166), and raised level of serum creatinine (35.84%; 119 out of 332).

There were 239 patients with abnormalities in their lung CT-scan, of whom 220 had different types of ground-glass opacities. Other forms of reported radiologic involvement were mixed pattern (118), reticular pattern (65), and honeycomb pattern (1). Involvements were more frequent in the lower lobes of both right and left lungs

From the total patients, 95 (26.84%) patients were Intubated and 28 (7.91%) supported by non-invasive ventilation whereas 64 (18.08%) of infected patients care without any support. The mean days of patient’s hospitalization were 5.98 ± 5.87 (± Standard deviation).

### Specific-Drug categories

Six sub-groups of patients were defined with respect to their drug histories. Each sub-group included patients who were under treatment with one of the following drugs or drug classes: aspirin, angiotensin II receptor blockers (ARBs), angiotensin-converting enzyme inhibitors (ACEIs), statins, clopidogrel, and metformin. The comprehensive information of these subgroups is presented in detail in Table 2.

**Table 2:** Baseline characteristics of COVID-19 patients, categorized by medication.

| Characteristics |  | Aspirin (%) | ACEIs (%) | ARBs (%) | Clopidogrel (%) | Metformin (%) | Statins (%) |
| --- | --- | --- | --- | --- | --- | --- | --- |
| Age | <50 | 8 (9.52) | 3 (21.43) | 7 (8.33) | 0 (0.00) | 5 (10.20) | 4 (6.45) |
|  | 50-65 | 28 (33.33) | 4 (28.57) | 23 (27.38) | 4 (21.05) | 22 (44.90) | 20 (32.26) |
|  | ≥65 | 48 (57.14) | 7 (50.00) | 54 (64.29) | 15 (78.95) | 22 (44.90) | 38 (61.29) |
| Gender | Female | 35 (41.67) | 5 (35.71) | 46 (54.76) | 7 (36.84) | 21 (42.86) | 27 (43.55) |
|  | Male | 49 (58.33) | 9 (64.29) | 38 (45.24) | 12 (63.16) | 28 (57.14) | 35 (56.45) |
| BMI | <35 | 59 (88.06) | 7 (87.50) | 63 (88.73) | 11 (91.67) | 34 (87.18) | 42 (87.50) |
|  | ≥35 | 8 (11.94) | 1 (12.50) | 8 (11.27) | 1 (8.33) | 5 (12.82) | 6 (12.50) |
| PMH | Diabetes | 43 (51.19) | 6 (42.86) | 42 (50.00) | 10 (52.63) | 48 (97.96) | 34 (54.84) |
|  | HTN | 52 (61.90) | 12 (85.71) | 75 (89.29) | 9 (47.37) | 26 (53.06) | 39 (62.90) |
|  | IHD | 54 (64.29) | 8 (57.14) | 32 (38.10) | 18 (94.74) | 15 (30.61) | 37 (59.68) |
|  | Organ transplant | 4 (4.76) | 0 (0.00) | 2 (2.38) | 0 (0.00) | 0 (0.00) | 1 (1.61) |
|  | HLP | 4 (4.76) | 2 (14.29) | 7 (8.33) | 1 (5.26) | 4 (8.16) | 11 (17.74) |
|  | Seizure | 0 (0.00) | 0 (0.00) | 2 (2.38) | 0 (0.00) | 0 (0.00) | 0 (0.00) |
|  | Thyroid disease | 2 (2.38) | 3 (21.43) | 4 (4.76) | 0 (0.00) | 3 (6.12) | 5 (8.06) |
|  | Cancer | 7 (8.33) | 2 (14.29) | 6 (7.14) | 2 (10.53) | 3 (6.12) | 4 (6.45) |
|  | AIDS | 0 (0.00) | 0 (0.00) | 0 (0.00) | 0 (0.00) | 0 (0.00) | 0 (0.00) |
|  | Immunodeficiency | 0 (0.00) | 0 (0.00) | 0 (0.00) | 0 (0.00) | 0 (0.00) | 0 (0.00) |
|  | ESRD | 3 (3.57) | 0 (0.00) | 2 (2.38) | 0 (0.00) | 0 (0.00) | 0 (0.00) |
|  | CVA | 5 (5.95) | 0 (0.00) | 4 (4.76) | 1 (5.26) | 0 (0.00) | 3 (4.84) |
|  | CKD | 6 (7.14) | 0 (0.00) | 6 (7.14) | 1 (5.26) | 1 (2.04) | 3 (4.84) |
| Symptom | Low Consciousness | 12 (14.29) | 0 (0.00) | 8 (9.52) | 2 (10.53) | 5 (10.20) | 6 (9.68) |
|  | Anorexia | 13 (15.48) | 1 (7.14) | 8 (9.52) | 1 (5.26) | 7 (14.29) | 8 (12.90) |
|  | No symptom | 3 (3.57) | 2 (14.29) | 1 (1.19) | 1 (5.26) | 1 (2.04) | 2 (3.23) |
|  | Cough | 39 (46.43) | 9 (64.29) | 41 (48.81) | 10 (52.63) | 20 (40.82) | 30 (48.39) |
|  | Dyspnea | 44 (52.38) | 9 (64.29) | 41 (48.81) | 14 (73.68) | 24 (48.98) | 33 (53.23) |
|  | Myalgia | 17 (20.24) | 1 (7.14) | 19 (22.62) | 2 (10.53) | 11 (22.45) | 12 (19.35) |
|  | Coryza | 0 (0.00) | 0 (0.00) | 1 (1.19) | 0 (0.00) | 1 (2.04) | 1 (1.61) |
|  | Nausea | 11 (13.10) | 1 (7.14) | 14 (16.67) | 1 (5.26) | 7 (14.29) | 10 (16.13) |
|  | Diarrhea | 6 (7.14) | 0 (0.00) | 4 (4.76) | 0 (0.00) | 4 (8.16) | 3 (4.84) |
|  | Headache | 2 (2.38) | 1 (7.14) | 2 (2.38) | 2 (10.53) | 1 (2.04) | 3 (4.84) |
|  | Chills | 8 (9.52) | 0 (0.00) | 6 (7.14) | 4 (21.05) | 6 (12.24) | 2 (3.23) |
|  | Sore throat | 4 (4.76) | 0 (0.00) | 5 (5.95) | 1 (5.26) | 4 (8.16) | 5 (8.06) |
|  | weakness | 39 (46.43) | 4 (28.57) | 35 (41.67) | 6 (31.58) | 19 (38.78) | 27 (43.55) |
|  | Anosmia | 0 (0.00) | 0 (0.00) | 1 (1.19) | 0 (0.00) | 0 (0.00) | 0 (0.00) |
|  | chest pain | 7 (8.33) | 0 (0.00) | 5 (5.95) | 2 (10.53) | 2 (4.08) | 2 (3.23) |
|  | others | 18 (21.43) | 3 (21.43) | 15 (17.86) | 5 (26.32) | 14 (28.57) | 15 (24.19) |
| Smoking |  |  |  |  |  |  |  |
|  | Non-smoker | 71 (84.52) | 12 (85.71) | 79 (94.05) | 15 (78.95) | 42 (85.71) | 52 (83.87) |
|  | Smoker | 7 (8.33) | 2 (14.29) | 2 (2.38) | 2 (10.53) | 6 (12.24) | 6 (9.68) |
|  | Ex-smoker | 6 (7.14) | 0 (0.00) | 3 (3.57) | 2 (10.53) | 1 (2.04) | 4 (6.45) |
| **Vital signs** |  |  |  |  |  |  |  |
| **O2 Saturation** | <90% | 11 (13.10) | 4 (28.57) | 17 (20.24) | 1 (5.26) | 6 (12.24) | 10 (16.13) |
|  | 90-96% | 39 (46.43) | 5 (35.71) | 36 (42.86) | 13 (68.42) | 21 (42.86) | 25 (40.32) |
|  | ≥96% | 34 (40.48) | 5 (35.71) | 31 (36.90) | 5 (26.32) | 22 (44.90) | 27 (43.55) |
| **Temperature** | <37.8 | 54 (64.29) | 11 (78.57) | 59 (70.24) | 14 (73.68) | 33 (67.35) | 41 (66.13) |
|  | 37.8- 39 | 24 (28.57) | 2 (14.29) | 21 (25.00) | 4 (21.05) | 15 (30.61) | 16 (25.81) |
|  | >39 | 6 (7.14) | 1 (7.14) | 4 (4.76) | 1 (5.26) | 1 (2.04) | 5 (8.06) |
| **Respiratory rate** | ≤25 | 75 (91.46) | 11 (78.57) | 78 (92.86) | 18 (94.74) | 44 (89.80) | 52 (85.25) |
|  | >25 | 7 (8.54) | 3 (21.43) | 6 (7.14) | 1 (5.26) | 5 (10.20) | 9 (14.75) |
| **Pulse rate** | ≤100 | 56 (67.47) | 11 (78.57) | 52 (61.90) | 13 (68.42) | 32 (65.31) | 42 (68.85) |
|  | >100 | 27 (32.53) | 3 (21.43) | 32 (38.10) | 6 (31.58) | 17 (34.69) | 19 (31.15) |
| **SBP** | 100-140 mmHg | 49 (58.33) | 7 (50.00) | 47 (55.95) | 12 (63.16) | 29 (59.18) | 36 (58.06) |
|  | <100 mmHg | 8 (9.52) | 0 (0.00) | 6 (7.14) | 2 (10.53) | 3 (6.12) | 3 (4.84) |
|  | >140 mmHg | 27 (32.14) | 7 (50.00) | 31 (36.90) | 5 (26.32) | 17 (34.69) | 23 (37.10) |
| **DBP** | 70-90 mmHg | 48 (57.14) | 4 (28.57) | 47 (55.95) | 10 (52.63) | 26 (53.06) | 38 (61.29) |
|  | <70 mmHg | 19 (22.62) | 4 (28.57) | 20 (23.81) | 6 (31.58) | 12 (24.49) | 13 (20.97) |
|  | >90 mmHg | 17 (20.24) | 6 (42.86) | 17 (20.24) | 3 (15.79) | 11 (22.45) | 11 (17.74) |
| **Laboratory data** |  |  |  |  |  |  |  |
| **Hb** | ≥10 | 71 (85.54) | 14 (100.00) | 66 (78.57) | 15 (83.33) | 39 (81.25) | 56 (90.32) |
|  | <10 | 12 (14.46) | 0 (0.00) | 18 (21.43) | 3 (16.67) | 8 (18.75) | 6 (9.68) |
| **PLT** | ≥150000 | 53 (63.86) | 12 (85.71) | 60 (71.43) | 16 (88.89) | 36 (75.00) | 42 (67.74) |
|  | <150000 | 30 (36.14) | 2 (14.29) | 24 (28.57) | 2 (11.11) | 12 (25.00) | 20 (32.26) |
| **WBC** | 4000-12000 | 55 (66.27) | 11 (78.57) | 54 (64.29) | 15 (83.33) | 37 (77.08) | 46 (74.19) |
|  | <4000 | 9 (10.84) | 2 (14.29) | 16 (19.05) | 0 (0.00) | 7 (14.58) | 5 (8.06) |
|  | >12000 | 19 (22.89) | 1 (7.14) | 14 (16.67) | 3 (16.67) | 4 (8.33) | 11 (17.74) |
| **Lymphocyte** | 1000-4000 | 31 (48.44) | 4 (40.00) | 32 (51.61) | 8 (61.54) | 19 (51.35) | 20 (43.48) |
|  | <1000 | 30 (46.88) | 6 (60.00) | 27 (43.55) | 5 (38.46) | 17 (45.95) | 25 (54.35) |
|  | >4000 | 3 (4.69) | 0 (0.00) | 3 (4.84) | 0 (0.00) | 1 (2.70) | 1 (2.17) |
| **Neutrophil** | 1500-7700 | 45 (70.31) | 9 (90.00) | 44 (70.97) | 9 (69.23) | 30 (81.08) | 33 (71.74) |
|  | <1500 | 0 (0.00) | 0 (0.00) | 2 (3.23) | 0 (0.00) | 2 (5.41) | 0 (0.00) |
|  | >7700 | 19 (29.69) | 1 (10.00) | 16 (25.81) | 4 (30.77) | 5 (13.51) | 13 (28.26) |
| **NLR** | 0 | 27 (61.36) | 3 (50.00) | 37 (74.00) | 8 (80.00) | 21 (80.77) | 18 (64.29) |
|  | 1 | 17 (38.64) | 3 (50.00) | 13 (26.00) | 2 (20.00) | 5 (19.23) | 10 (35.71) |
| **CRP** | Negative | 37 (46.25) | 7 (58.33) | 47 (58.02) | 6 (33.33) | 26 (55.32) | 32 (53.33) |
|  | Positive | 43 (53.75) | 5 (41.67) | 34 (41.98) | 12 (66.67) | 21 (44.68) | 28 (46.67) |
| **AST** | ≤40 | 36 (50.70) | 8 (61.54) | 41 (59.42) | 7 (46.67) | 14 (35.00) | 25 (48.08) |
|  | >40 | 35 (49.30) | 5 (38.46) | 28 (40.58) | 8 (53.33) | 26 (65.00) | 27 (51.92) |
| **ALT** | ≤40 | 54 (73.97) | 8 (61.54) | 54 (77.14) | 11 (73.33) | 29 (72.50) | 37 (69.81) |
|  | >40 | 19 (26.03) | 5 (38.46) | 16 (22.86) | 4 (26.67) | 11 (27.50) | 16 (30.19) |
| **ALK-p** | ≤200 | 41 (58.57) | 4 (30.77) | 45 (67.16) | 6 (42.86) | 24 (61.54) | 30 (57.69) |
|  | >200 | 29 (41.43) | 9 (69.23) | 22 (32.84) | 8 (57.14) | 15 (38.46) | 22 (42.31) |
| **Total Bill** | ≤1.5 | 45 (80.36) | 9 (100.00) | 42 (80.77) | 9 (90.00) | 20 (76.92) | 35 (76.09) |
|  | >1.5 | 11 (19.64) | 0 (0.00) | 10 (19.23) | 1 (10.00) | 6 (23.08) | 11 (23.91) |
| **PTT** | ≤45 | 65 (94.20) | 10 (83.33) | 63 (94.03) | 13 (92.86) | 38 (100.00) | 47 (95.92) |
|  | >45 | 4 (5.80) | 2 (16.67) | 4 (5.97) | 1 (7.14) | 0 (0.00) | 2 (4.08) |
| **INR** | ≤1.5 | 54 (76.06) | 10 (83.33) | 60 (83.33) | 12 (80.00) | 34 (85.00) | 38 (76.00) |
|  | >1.5 | 17 (23.94) | 2 (16.67) | 12 (16.67) | 3 (20.00) | 6 (15.00) | 12 (24.00) |
| **PH** | 7.35-7.45 | 56 (67.47) | 11 (78.57) | 56 (67.47) | 13 (72.22) | 37(77.08) | 40 (64.52) |
|  | <7.25 | 8 (9.64) | 1 (7.14) | 5 (6.02) | 0 (0.00) | 1 (2.08) | 5 (8.06) |
|  | 7.25-7.35 | 11 (13.25) | 1 (7.14) | 10 (12.05) | 2 (11.11) | 6 (12.50) | 9 (14.52) |
|  | >7.45 | 8 (9.64) | 1 (7.14) | 12 (14.46) | 3 (16.67) | 4 (8.33) | 8 (12.90) |
| **PCo2** | 35-45 | 27 (33.75) | 5 (41.67) | 31 (40.26) | 5 (29.41) | 19 (39.58) | 22 (36.07) |
|  | <35 | 28 (35.00) | 3 (25.00) | 27 (35.06) | 9 (52.94) | 15 (31.25) | 18 (29.51) |
|  | >40 | 25 (31.25) | 4 (33.33) | 19 (24.68) | 3 (17.65) | 14 (29.17) | 21 (34.43) |
| **Hco3** | <22 | 31 (38.75) | 5 (41.67) | 31 (40.26) | 7 (41.18) | 16 (33.33) | 26 (42.62) |
|  | 22-26 | 27 (33.75) | 4 (33.33) | 25 (32.47) | 7 (41.18) | 11 (22.92) | 16 (26.23) |
|  | >26 | 22 (27.50) | 3 (25.00) | 21 (27.27) | 3 (17.65) | 21 (43.75) | 19 (31.15) |
| **K** | 3.5-5 | 63 (75.90) | 12 (92.31) | 61 (73.49) | 14 (77.78) | 43 (91.49) | 50 (80.65) |
|  | <3.5 | 0 (0.00) | 0 (0.00) | 3 (3.61) | 0 (0.00) | 0 (0.00) | 0 (0.00) |
|  | >5 | 20 (24.10) | 1 (7.69) | 19 (22.89) | 4 (22.22) | 4 (8.51) | 12 (19.88) |
| **Na** | 135-145 | 69 (83.13) | 13 (100.00) | 69 (83.13) | 13 (72.22) | 38 (80.85) | 49 (79.03) |
|  | <135 | 10 (12.05) | 0 (0.00) | 10 (12.05) | 3 (16.67) | 6 (12.77) | 9 (14.52) |
|  | >145 | 4 (4.82) | 0 (0.00) | 4 (4.82) | 2 (11.11) | 3 (6.38) | 4 (6.45) |
| **Cr** | ≤1.2 | 46 (55.42) | 6 (42.86) | 44 (52.38) | 6 (33.33) | 31 (64.58) | 34 (54.84) |
|  | >1.2 | 37 (44.58) | 8 (57.14) | 40 (47.62) | 12 (66.67) | 17 (35.42) | 28 (45.16) |
| **U/A** | hematuria | 21 (48.84) | 2 (28.57) | 16 (35.56) | 3 (30.00) | 6 (30.00) | 11 (35.48) |
|  | leukocyturia | 8 (18.60) | 2 (28.57) | 5 (11.11) | 2 (20.00) | 5 (25.00) | 5 (16.13) |
|  | proteinuria | 22 (51.16) | 3 (42.86) | 16 (35.56) | 5 (50.00) | 9 (45.00) | 13 (41.94) |
| **COVID PCR** | Positive | 46 (58.23) | 3 (25.00) | 49 (62.03) | 7 (38.89) | 24 (54.55) | 30 (51.72) |
| **Radiology findings** |  |  |  |  |  |  |  |
|  | ground-glass opacity | 61 (95.31) | 6 (66.67) | 51 (86.44) | 10 (90.91) | 32 (91.43) | 37 (92.50) |
|  | consolidation | 49 (76.56) | 9 (100.00) | 48 (81.36) | 9 (81.82) | 27 (77.14) | 27 (67.50) |
|  | bilateral involvement | 60 (93.75) | 8 (88.89) | 55 (93.22) | 11 (100.00) | 34 (97.14) | 37 (92.50) |

ACEIs: Angiotensin-converting enzyme inhibitors; ARBs: angiotensin II receptor blockers; BMI: Body mass index; HTN: Hypertension; IHD: Ischemic heart disease; HLP: Hyperlipidaemia; AIDS: Acquired immunodeficiency syndrome; ESRD: End stage renal disease; CVA: Cerebrovascular Accident; CKD: Chronic kidney disease; SBP: Systolic blood pressure; DBP: Diastolic blood pressure; HB: Haemoglobin; PLT: Platelets; WBC: White blood cell; NLR: Neutrophil to lymphocyte ratio; CRP: C-reactive protein; AST: Aspartate aminotransferase; ALT: Alanine aminotransferase; ALK-p: alkaline phosphatase; Bill: bilirubin; U/A: Urine analysis.

We explored data in our registry to find notable correlations of drug use with different aspects of COVID-19. With regards to the clinical manifestations of the disease it is noteworthy that, as our data suggests, previous use of aspirin is correlated with decreased level of consciousness upon admission (p < 0.05), and on the other hand, patients who were under treatment with ACEIs are more probable to be symptom-free (p < 0.05).

Radiologic involvement of the lungs showed a quite specific pattern with respect to the drug used (Table 3). Ground-glass opacities were more common in those receiving ARBs (p < 0.05) and ACEI (p < 0.05), consolidations with statins (p < 0.05), and emphysema and its severity with clopidogrel (p < 0.05) and metformin (p = 0.005) respectively. Chest lymphadenopathies were more probable in the setting of using aspirin (p < 0.005), Clopidogrel (p < 0.005), and ACEI (p < 0.05). Specific localization pattern of lung involvement was concomitant with the usage of metformin and ACEI. Metformin usage is correlated with the involvement of left lung lower lobe (p < 0.05), whereas ACEI usage is associated with right lung involvements, revealed by its correlation with the upper lobe involvement (p < 0.05) and lower lobe involvement score (p < 0.05).

**Table 3:** Radiology involvement score in COVID-19 patients in different classes of drug use.

|  | Aspirin | ACEIs | ARBs | Clopidogrel | Metformin | Statins |
| --- | --- | --- | --- | --- | --- | --- |
| **Radiology involvement score correlation** | 0.5625 | 0.5678 | 0.5919 | 0.3696 | 0.9725 | 0.4772 |

Correlations between the pattern of laboratory results and drug usage were detected in our study. Notable findings were the association of aspirin with elevated levels of d-dimer (p < 0.05), ARBs with the elevated levels of cardiac troponin I (CTNI) (p < 0.05), lactate dehydrogenase (LDH) (p < 0.05) and lactate (p < 0.05), ACEI with the elevated level of LDH (p < 0.05), metformin with high levels of CTNI (p < 0.005) and amylase (p < 0.05), and Clopidogrel with high levels of amylase (p = 0.005).

After conducting the analysis, the clinical data of about 100 other patients were added to our database. We re-evaluated our data and re-analysed them. The results were totally consistent with what is presented here and no significant change was found to exclude any of the previous results. However, it is noteworthy that these new analyses suggested that there is a correlation between the usage of statins and aspirin with having a normal white blood cell and lymphocyte count upon admission. Results are not shown.

The outcome of patients, including mortality, ventilation requirement and ICU admission showed no correlation with receiving ARBs, ACEIs, statins, or Clopidogrel. However, aspirin-users were more likely to need no ventilation support (p = 0.05), whereas, metformin-usage were more associated with the chance of intubation in the course of hospitalization (p < 0.05) and also of the mortality (p < 0.05). Details are shown in Table 4.

**Table 4:**
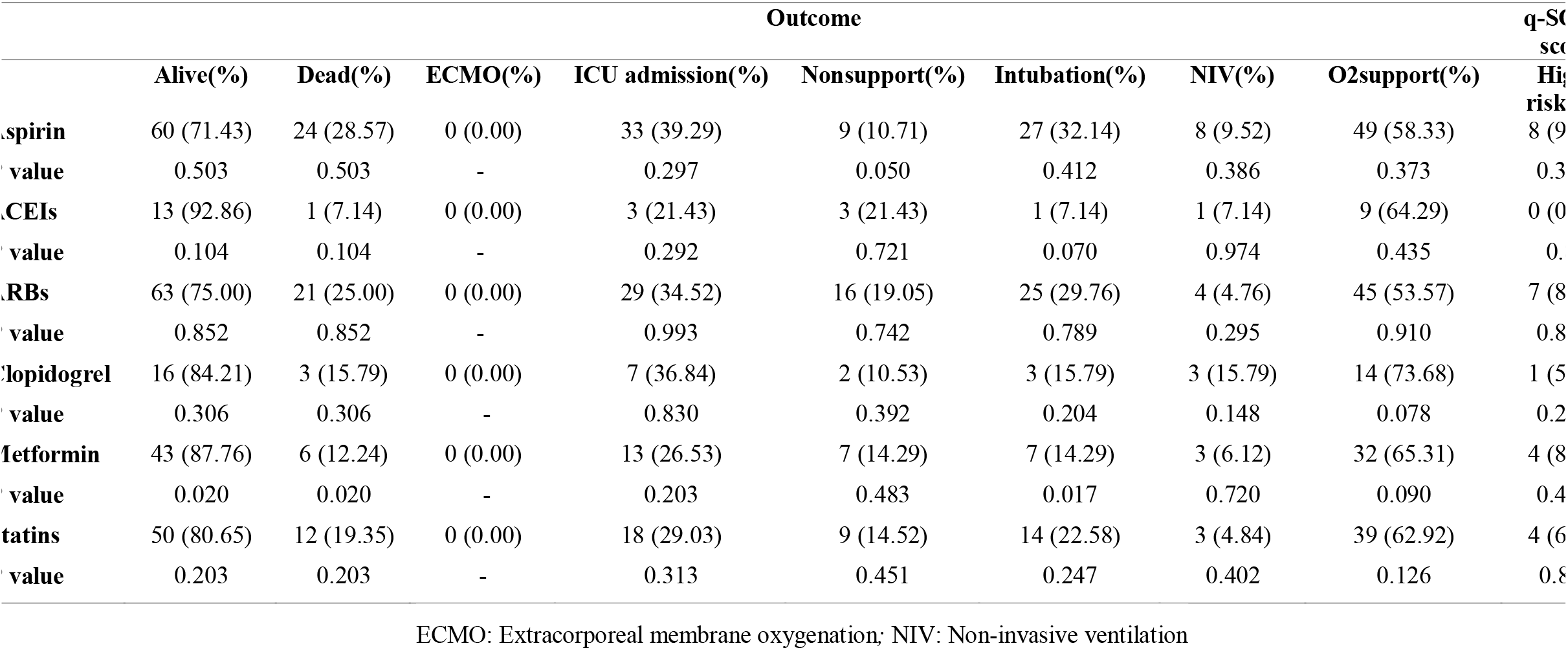
Outcomes of patients with COVID-19 with regards to previous medication use.

## Discussion

There are many studies showing positive and negative effects of drugs like statins, ACEI, ARBS, metformin. and anti-platelets. We have shown that COVID-19 patients receiving these drugs prior to admission were neither better nor worse regarding their outcomes. In theory, there are at least 4 reasons statins might be useful for COVID-19 patients. First, cardiovascular disease is the most significant risk factor (10-15% case fatality rate)[10] for severe COVID-19, therefore, these patients would likely already benefit from their use. Second, a number of cardiovascular complications such as thrombosis and myocarditis have been reported in association with this disease and statins might be beneficial in preventing them. Third, statins might have a role in protecting innate immune responses to viral respiratory infections (including to SARS-CoV-2) via inhibiting the MYD88 pathway. Usually, statins do not change the level of MYD88, they only keep its level in a normal range during hypoxia and stress [11]. And last but not least, epidemiological studies have shown that statins may prevent severe viral pneumonias [12].

A study conducted in Belgium reported that nursing home residents taking a statin were three times more likely to be free of symptoms of COVID-19 during their infection than those who did not. Length of admission and death was also slightly less in those receiving statins, even though the difference was not significant statistically [13].

We have shown that statins did not protect COVID-19 patients from having worse outcomes, although those receiving statins probably had baseline cardiovascular diseases and/or diabetes which could have worsened their prognosis per se. Statins have been widely prescribed with a good safety index.

Data on anti-platelets such as aspirin and clopidogrel and their effects on COVID-19 are scarce. We didn’t find any significant difference in outcomes in our patients. However, thrombosis remains a major complication of this disease and trails on anticoagulants are showing promise.

Zhang P. et al. conducted a retrospective, multicentre study of 1,128 COVID-19 patients with hypertension in which 188 were taking ACEI/ARBs. They concluded that these patients had a lower all-cause mortality compared to those not receiving ACEI/ARBs [14].

Physiological models of SARS-CoV infection have shown a theoretical benefit of ACEI/ARBs, however, this benefit cannot be attributed to SARS-CoV-2. Cardiology associations such as the ACC, HFSA, AHA, and ESC Hypertension Council, have rejected this hypothesis. A commentary published in the Lancet Respiratory Medicine even proposed that ACEI might increase the risk for severe COVID-19 infection [15].

ACE2 receptor upregulation results in increased binding sites for SARS-CoV-2, leading to a higher risk for COVID-19 infection. Ferrario et al. showed that Lisinopril and losartan caused a 5 and 3-fold increase in ACE2 levels, respectively [16].

Li et al. hypothesized that ACEI could stimulate a negative feedback [4], while Sun et al. argued that their use could lead to a decreased chance of SARS-CoV-2 entering the cell [17].

The American College of Cardiology and American Heart Association (ACC/AHA) states that “there are no experimental or clinical data demonstrating beneficial or adverse outcomes with background use of ACE inhibitors or ARBs.” This statement recommends continuing these drugs if they are being prescribed for valid indications and advises clinicians not to add or remove them “beyond actions based on standard clinical practice. Available at: (https://viajwat.ch/2REZU2H)

Given the common use of ACE inhibitors and ARBs worldwide and reviewing the literature along with our study, we provide tentative reassurance that at least ACEI and ARBs are safe in patients with COVID-19. Whether they are actually beneficial should be studied in clinical trials. There is an urgent need for available and safe drugs to treat COVID-19, however, we must take caution in their prescription since they may exacerbate this disease. We recommend guideline-directed administration and continuation of these drugs, and we emphasize that our primary role as physicians is to do no harm.

## Conclusions

Our Findings could be assumed to be suggestive for certain correlations between the presentations and the course of COVID-19 infection, and the distinct drug groups used before and during the course of the disease by the patients. However, our specific positive results were not in accord with the findings of previously done studies. Moreover, we could not replicate the positive findings, i.e. the specific correlations between medical conditions and drug usages, of previous studies as it is discussed in details in the previous section. Therefore, it is reasonable to conclude that further investigations are crucial to bring about the conclusive evidences with regards to the effects of prior usage of aspirin, statins, metformin, ARBs, ACEIs, and Clopidogrel on the outcome and various presentations of COVID-19.

Summary Points
- **From the total of 353 patients, 57.51% cases were male. Patients older than 65- years accounted for 45% of all cases**.
- **With regards to the past medical history, 129 (36.54%) patients were found to have hypertension and 111 (31.44%) had diabetes**.
- **The most prevalent symptoms were as following: 198 (56.09%) dyspnea, 178 (50.42%) fever, and 177 (50.14%) coughing**.
- **Association of aspirin with elevated levels of d-dimer (p < 0.05), ARBs with the elevated levels of cardiac troponin I (CTNI) (p < 0.05), lactate dehydrogenase (LDH) (p < 0.05) and lactate (p < 0.05), ACEI with the elevated level of LDH (p < 0.05), metformin with high levels of CTNI (p < 0.005) and amylase (p < 0.05), and Clopidogrel with high levels of amylase (p = 0.005) were the remarkable findings of laboratory data**.
- **There were 239 patients with abnormalities in their lung CT-scan. The ground-glass opacities were the most frequent form of involvements**.
- **The outcome of patients, including mortality, ventilation requirement and ICU admission showed no correlation with receiving ARBs, ACEIs, statins, or Clopidogrel**.
- **Aspirin-users were more likely to need no ventilation support (p = 0.05), whereas, metformin-usage were more associated with the chance of intubation in the course of hospitalization (p < 0.05) and also of the mortality (p < 0.05)**.
- **Patients who were under treatment with ACEIs are more probable to be symptom-free (p < 0.05)**.

## Data Availability

We shall declare that we are not allowed to disclose publically our data. Individual researchers and research institutes can request to access data by contacting the corresponding author.

## Author contributions

AA and MS designed the study; AK performed statistical analysis; AR analyzed the radiological documents; AA and NS wrote and revised the manuscript. BK, AA, AS, ME, AS, SMP, MS, MA all collaborated in gathering data and confirmed the final version of manuscript.

## Acknowledgements

We thank all our patients who participated in the study.

## Financial disclosure

The authors have no relevant affiliations or financial involvement with any organization or entity with a financial interest in or financial conflict with the subject matter or materials discussed in the manuscript. This includes employment, consultancies, honoraria, stock ownership or options, expert testimony, grants or patents received or pending, or royalties.

No writing assistance was utilized in the production of this manuscript.

## Ethics disclosure

The study was approved by the ethics committee of Tehran University of Medical Sciences. The ethics committee waived the requirement for informed patient consent for this retrospective study subject to the anonymity of patients.

Papers of special interest are highlighted as: •

